# Two-Step Light Gradient Boosted Model to identify human West Nile Virus infection risk factor in Chicago

**DOI:** 10.1101/2023.05.09.23289737

**Authors:** Guangya (Wayne) Wan, Joshua Allen, Weihao Ge, Shubham Rawlani, John Uelmen, Liudmila Sergeevna Mainzer, Rebecca Lee Smith

## Abstract

West Nile virus (WNV), a flavivirus transmitted by mosquito bites, causes primarily mild symptoms but can also be fatal. Therefore, predicting and controlling the spread of West Nile virus is essential for public health in endemic areas. We hypothesized that socioeconomic factors may influence human risk from WNV. We analyzed a list of weather, land use, mosquito surveillance, and socioeconomic variables for predicting WNV cases in 1-km hexagonal grids across the Chicago metropolitan area. We used a two-stage lightGBM approach to perform the analysis and found that hexagons with incomes above and below the median are influenced by the same top characteristics. We found that weather factors and mosquito infection rates were the strongest common factors. Land use and socioeconomic variables had relatively small contributions in predicting WNV cases. The Light GBM handles unbalanced data sets well and provides meaningful predictions of the risk of epidemic disease outbreaks.

## Introduction

West Nile Virus (WNV) is a mosquito-borne flavivirus that has been circulating in the United States for two decades, first appearing in New York in 1999 [1–3]. The disease is spread in an enzootic mosquito-bird-mosquito circulation [4–7], and zoonotic transmission occurs when humans are bitten by a WNV-positive mosquito [8]. Because there are no vaccines for WNV in humans, prediction of WNV-positive mosquitoes is used to inform public health actions to clear mosquitoes in areas of high risk [9] and to warn the general public of increased risk.

Efforts have been made to build predictive models of WNV spread [10]. Predicting human cases would help to identify high-risk populations, and therefore enable protective measures. Paz [11] analyzed major weather factors and found temperature and precipitation are associated with WNV human cases. A temperature range of 10-35°C is advantageous for mosquito breeding activity. However, an association of temperature with WNV infection risk is not always positive. Hahn et al.[12] performed a climate-region-wise analysis and found that in most regions of the US, temperature above the local average increases WNV risk, while in the western regions of the US, above-average temperature decreases WNV risk. Shocket et al. [13] has identified the optimal temperature range for mosquitoes that vector WNV is between 23-26°C. Precipitation and humidity have complex associations with mosquito population and infection rate, as well. Interaction between temperature and precipitation also explains a significant part of the WNV mosquito infection rate [14]. Poh et al. identified that temperature and rainfall increase mosquito abundance [15]. In addition to temperature and precipitation, other factors such as humidity and wind velocity affect mosquito abundance [16]. Peper et al. have studied WNV and mosquito surveillance records from Lubbock, TX, and have found that the probability of mosquito infection depends on the weather variables including the time in the year, wind, visibility, humidity, dew point, and the time lag of these variables [17]. They also found that weather has a temporal autocorrelation, which brings lagging effects into play [18,19]. DeFelice has discussed the lag in reporting of both mosquito infection and human cases that reduces real-time WNV forecast accuracy and proposed recursive optimization and Poisson process simulation for the retrospective forecast to solve the problem [20]. The landscape also contributes to WNV risk. Studies have identified geological factors such as vegetation, urbanization, mosquito breeding sites, and wetlands to be associated with WNV incidences [21–23]. Sánchez-Gómez et al. have discussed how temperature and the presence of wetlands influence WNV circulation in vectors and humans [21]. Hernandez et al. have identified weather, demographic, and controlling measurements including temperature, precipitation, ethnicity, mosquito breeding sites, targeted prevention, and education as key predictors [22]. Myer and Johnston have analyzed a 15-year span of data in Nassau County, NY, and identified landscape factors including high normalized difference vegetation index (NDVI), wetlands, and high urban development have a negative association with WNV incidences [23]. Farooq et.al. have estimated WNV expansion risk and found early spring weather, population, and agriculture activities can be important factors for early warning systems to predict Europe WNV outbreak [24]. Bassal et.al. investigated demographic disparities for WNV IgG levels in Israel and identified different WNV seroprevalence among geographical regions. Bassal et. al. also discovered different prevalence among racial groups, which have different socioeconomic status [25].

Linear regression and ensemble tree methods are the two most commonly used approaches for predicting WNV incidence or mosquito populations. Hernandez et al. started with chi-squared tests to identify a list of candidate factors and then used regression to find the strongest predictors [22]. Karki et al. used a stepwise model selection procedure to automatically test all factors and find the strongest predictors [26]. However, the risk of WNV is not linear with the factors. Furthermore, linear models have high specificity and perform best when there are no cases of viral infection, but have poor sensitivity when there are cases (low recall). To address these two issues, ensemble methods, specifically light gradient boosting method (GBM) approach [27], are used as our model in this paper. Light GBM is based on building an ensemble of decision trees instead of a single model to make the prediction. Therefore, neither requires linearity in the problem. However, light GBM is much faster to train and evaluate than other methods such as random forest [28,29], has a generally lower bias, and thus will be our focus in this paper. We performed a two-step light GBM approach as recommended for other ensemble tree methods [28,29]. In the first step, all factors are included in the model. And then a second light GBM classification/regression is performed based on the top factors selected by the first model [28].

We have hypothesized that, in addition to natural factors such as mosquito infection rate (MIR), weekly temperature, temperature in January, and precipitation, social economics and land cover factors will also be predictive factors for the WNV occurrences. We also hypothesized that natural factors might have lagging effects. These effects, linear or not, can be detected by the light GBM approach and identify areas at high risk of WNV cases and provide guidance for health intervention.

## Methods

### Data Set and pre-analysis

The dataset we used is described in more detail in Karki, et al. [26]. The dataset includes the number of human disease cases from 2005-2016 in Cook and DuPage Counties, IL, as the dependent variable, and several independent variables comprising weather, socioeconomic, land cover, and mosquito infection rates (MIR). All variables were aggregated on a weekly temporal resolution and on a spatial grid of 1 km wide hexagons for the study region.

The human disease data is described as a binary number that represents whether a case occurs in a hexagon in a given week. We performed the two-sample Kolmogorov-Smirnov (KS) test [30] and the two-step light GBM classification [27] to build the model to predict the human illness data and to derive the illness probability from the model.

Weather variables include temperature and precipitation, as well as the lagged variables representing temperature and precipitation 1 week, 2 weeks, 3 weeks, and 4 weeks before human case report date. The original weather data was collected by PRISM [31], aggregated to census tract level, and mapped to hexagons by Karki, et al. [26]

The land cover data include urban areas (developed open space, developed low intensity, developed medium intensity, developed high intensity), forest (deciduous, evergreen, and mixed), barren land, shrubs, grassland, pasture, cultivated crops, woody wetlands, herbaceous wetlands, and open water. Karki, et al.[26] retrieved the land cover data from the 2016 National Land Cover Database (NLCD) [32] and aggregated the percentage of different land covers in the hexagons.

For the socioeconomic data used by Karki, et al. [26], the 2016 census data from the US Census Bureau[33] was applied across all years. The data were converted from the census tract level to the hexagon level by assuming homogeneous socioeconomic status within each census tract. To determine the sensitivity of the socioeconomic data to annual changes, we replicated the mapping procedure with the 5-year rolling averages from 2010-2017 and performed the model analysis with both datasets (S1, S2). We found that the results are similar and the conclusions do not change; therefore, we will present the model built with the 2016 census data.

The variables we used are listed in Table 1 below.

**Table 1.** List of variables involved in building the models. We have variables representing nature factors, land cover, and socioeconomic data.

| Notation | Variable | Type |
| --- | --- | --- |
| mir_mean,<br>mir_lag1-4 | Mosquito Infection Rate (MIR) measured 0-4 weeks before human case report date | nature |
| preci,<br>preci_lag1-4 | Precipitation measured 0-4 weeks before human case report date |  |
| tempc,<br>temp_lag1-4 | Temperature measured 0-4 weeks before human case report date |  |
| tempJan | Temperature in January |  |
| dospct | Proportion of developed open space | Land cover |
| dlipect | Proportion of developed low intensity |  |
| dmipct | Proportion of developed medium intensity |  |
| dhipct | Proportion of developed high intensity |  |
| dfpct | Proportion of deciduous forests |  |
| efpct | Proportion of evergreen forests |  |
| mfpct | Proportion of mixed forests |  |
| blpct | Proportion of barren land |  |
| shrubspct | Proportion of shrubs |  |
| glandpct | Proportion of grassland |  |
| pasturepct | Proportion of pasture |  |
| clpct | Proportion of cultivated land |  |
| wwpct | Proportion of woody wetlands |  |
| shwpct | Proportion of herbaceous wetland |  |
| owpct | Proportion of open water |  |
| hpctpreww,<br>hpctpostww,<br>hpct7089,<br>hpctpost90 | Percentage of houses built before WWII, after WWII,<br>between 1970-1989, and after 1990 |  |
| whitepct,<br>blackpct,<br>asianpct,<br>hispanicpct | Percentage of white, african american, asian, and hispanic<br>population | Demographic,<br>socioeconomics |
| tot_pop | Total population |  |
| Income | Median household income |  |

We performed the two-sample Kolmogorov-Smirnov (KS) test [30] as a univariate analysis to identify the candidate key predictors to get some insights that would be helpful before building the models. The KS test is a model-free approach to test whether the distributions of features corresponding to two different classes behave similarly. Therefore, we did not reject collinearity in the KS test. A list of p-values was calculated to assess the importance of the features. We examined the distributions of the weather, socioeconomic, and land cover factors separately for the presence and absence of human cases by hexagon and week. The KS test would indicate which variables are distributed differently for the two situations. The KS score will serve as a criteria in choosing which variables to keep among a set of highly-correlated variables. For each set of highly-correlated variables, we will keep the one with the highest KS score.

Before building the light GBM model, we first assessed collinearity by generating the covariance plots calculated from Pearson’s correlation. We set the correlation threshold at 0.35 and kept the variables with the largest −log(p) values in the KS test. Therefore, it is possible that the factors selected in the model are correlated with the true predictors. We then selected the variables with the highest functional significance to build the models. We also evaluated the income-stratified data to check whether the high-income and low-income groups have different characteristics (S3).

### Two-step Light GBM Modeling

The hyperparameter for the light GBM is tuned with grid search with a predefined set, evaluated on the metric log-loss score as the decision criterion, which can help deal with the highly zero-inflated characteristic of the WNV case number. We used the lightgbm package in Python [34] to perform the light GBM method.

The model was built using a heuristic approach with two light GBM categorization procedures. After removing the correlation, we ran the first light GBM procedure on all remaining variables. We then examined the distribution of feature importance, selected the top variables by the natural gap in the distribution, and ran another light GBM procedure. Feature importance is defined as the mean decrease in impurity when a given feature is included to split the WNV_binary = 0 and WNV_binary = 1 cases. Feature importance is represented by the negative logarithm of the absolute value of importance. We evaluated the receiver operating characteristics area under the curve (ROC-AUC) to find the best threshold for a minimum model. The ROC-AUC score is insensitive to imbalanced data. With the threshold identified, we are able to evaluate the accuracy, recall, precision, and F-1 score [35]. We first fit the model with high and low income data to confirm that the models are similar (S3). Therefore, we build our model based on the full dataset. We then examine the distribution of feature importance and select subsets of features to build reduced models. We examine the performance of the reduced models to find a minimal model that retains predictive power.

Then, in the final model, we evaluated the relative importance of the covariates to identify important predictive features for WNV cases in our models. For the features of interest, we generate partial dependence (PD) plots to show their marginal predicted probability. The slope of the PD plot represents the strength of the feature. The shape of the PD plot could also indicate whether the effect is monotonic. The PD plots could easily show the nonlinear effects that are difficult to identify by regression.

## Results

### KS test

We performed univariable KS tests on all variables (Figure 1). We found that temperatures and mosquito infection rates have significant effects in the model. On the other hand, precipitation, land cover and socio-economic characteristics do not contribute significantly to the WNV risk.

**Fig 1.**
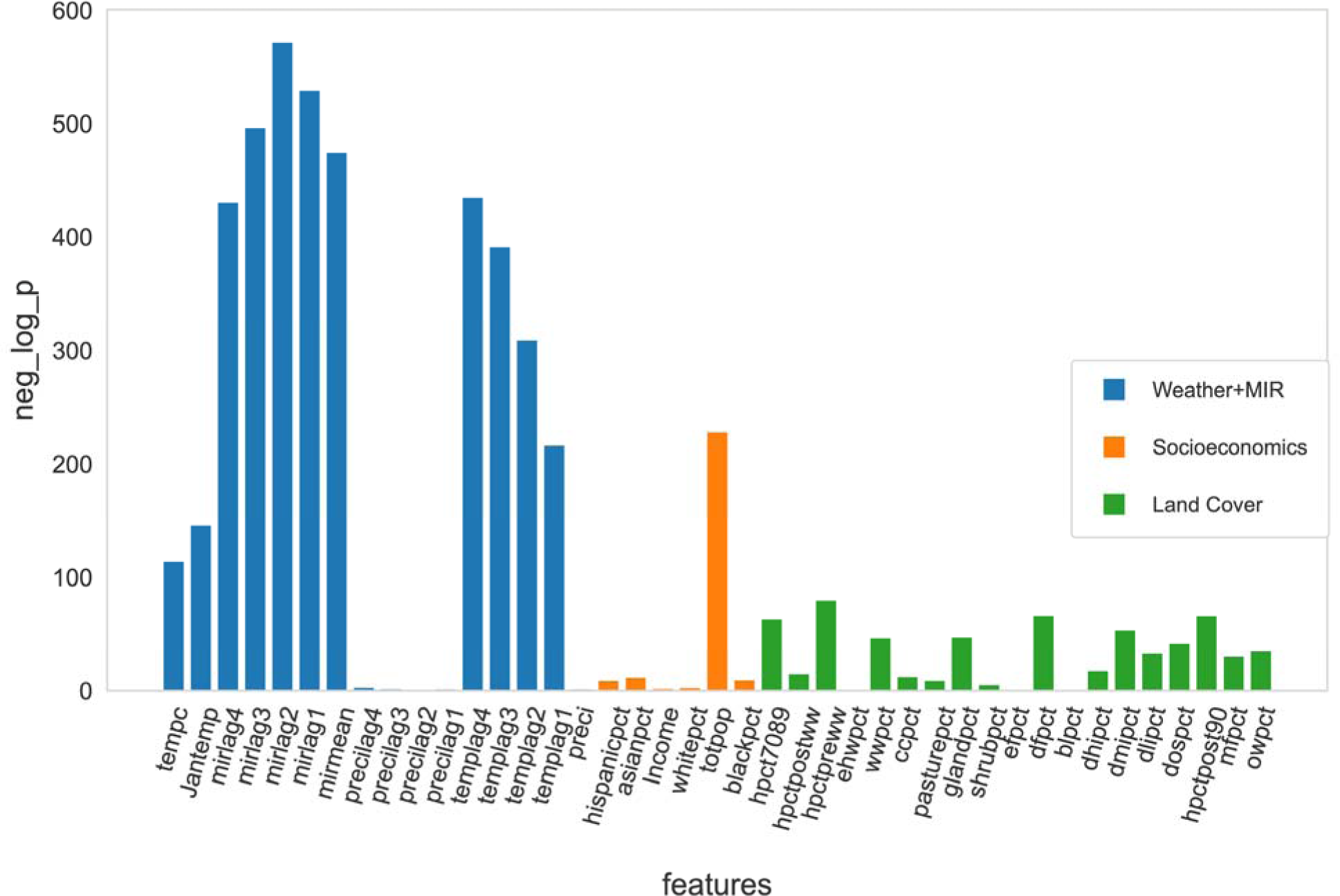
−log(p) of Kolmogorov-Smirnov test for all the features and covariates. From the KS test, we calculate the p-value, which indicates how different the distribution of the variable is between the hexagon-weeks with and without a case. The larger the −log(p), the less similar the two distributions are. The variables are grouped into four main categories + one residue category, but we have combined the fast-changing weather and MIR into the same category because these variables, as well as the number of cases, are captured with a temporal resolution of one week. Green bars represent land cover variables. Orange bars represent socio-economic variables. The blue bars represent strongly fluctuating variables: weather and mosquito infection rate.

### Variable correlations

Figure 2 shows the correlation between the variables. We found that weekly temperatures have a strong positive temporal correlation (0.47 - 0.84). On the other hand, the lagged effects of weekly MIR (0.075 - 0.18) and weekly precipitation (−0.022 - 0.044) are not as strongly correlated. Weekly MIR and weekly precipitation are also independent of other variables.

**Fig 2.**
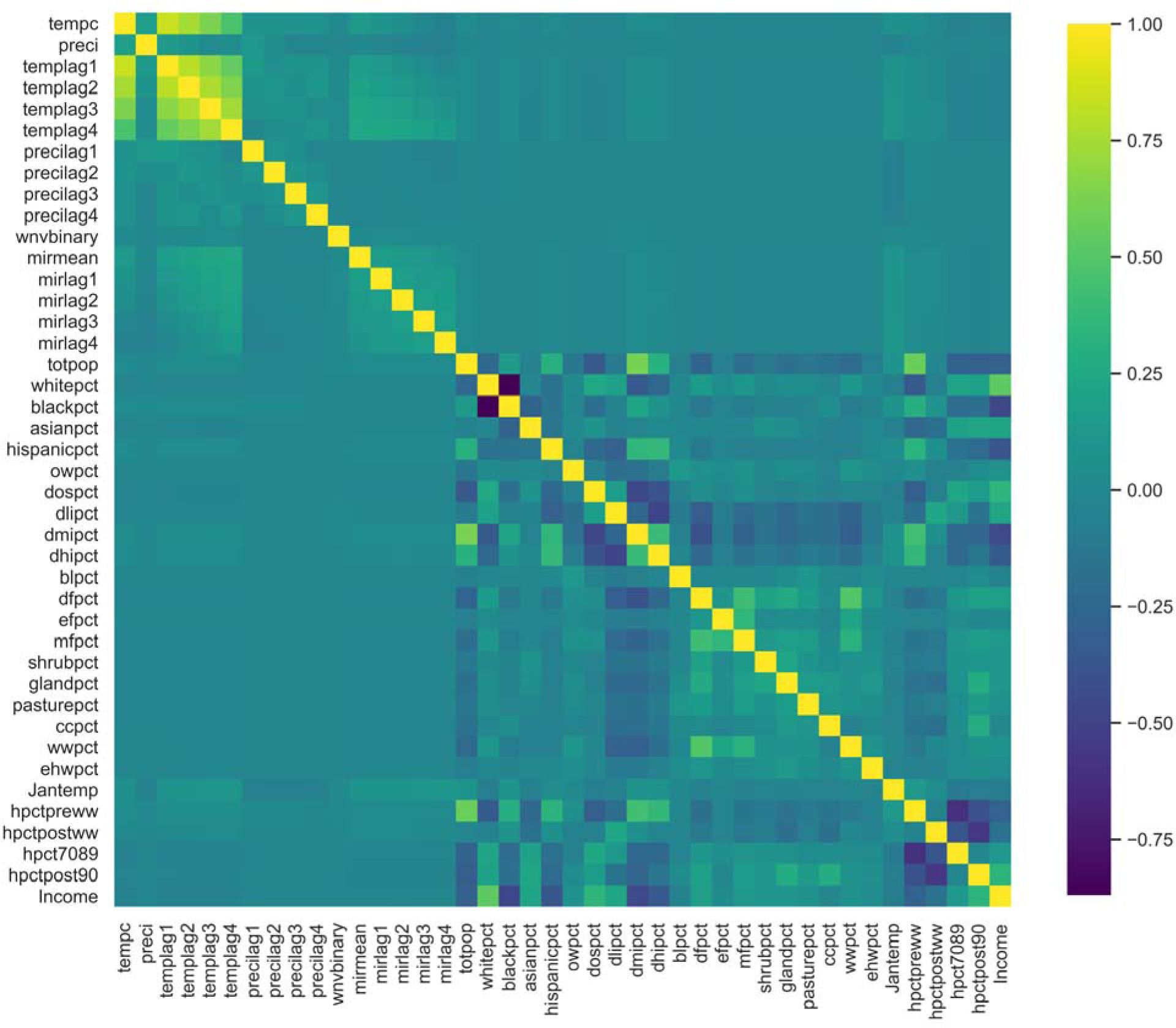
Heat-Map Covariance matrix for all the features. Original data are from Karki (2020) [26]. Yellow colors indicate strong positive correlations; dark blue colors indicate strong negative correlations. Light blue or green colors indicate weak correlations. We infer that temperature has a relatively high temporal correlation, as the variables tempc and templag1-4 (current temperature and temperatures 1-4 weeks before) are correlated. In addition, development stage and housing age are correlated with population, showing the interaction of population aggregation with land cover and housing status.

We also found that income is strongly correlated with race. Income has a high positive correlation (0.54) with the white race percentage in the hexagon area, and a medium-high negative correlation with the black race percentage (−0.46) and the Hispanic race percentage (−0.37). The white and black population percentages have a strong negative correlation with each other (−0.87), which is to be expected since the total population percentages should add up to 100%.

For each set of medium to highly correlated variables, we kept the variables with the highest KS scores for the light GBM analysis. The remaining variables are: All precipitation and MIR variables, mean temperature of 4 weeks before the human case report, mean temperature in January, total population, proportion of developed low intensity, proportion of open water, proportion of barren land, proportion of evergreen forest, proportion of shrubs, proportion of grassland, proportion of pasture, proportion of cultivated land, proportion of woody wetlands, emergent herbaceous wetlands, percent temperature in January, house post World War II, and income.

### Light GBM based on all selected features

We built our models using cross-validation, randomly splitting training and test sets, and then selected the best parameter based on the log-loss criteria. The importance of each predictor in the model is shown in Figure 3, and its performance on the test set is shown in Table 2.

**Fig 3:**
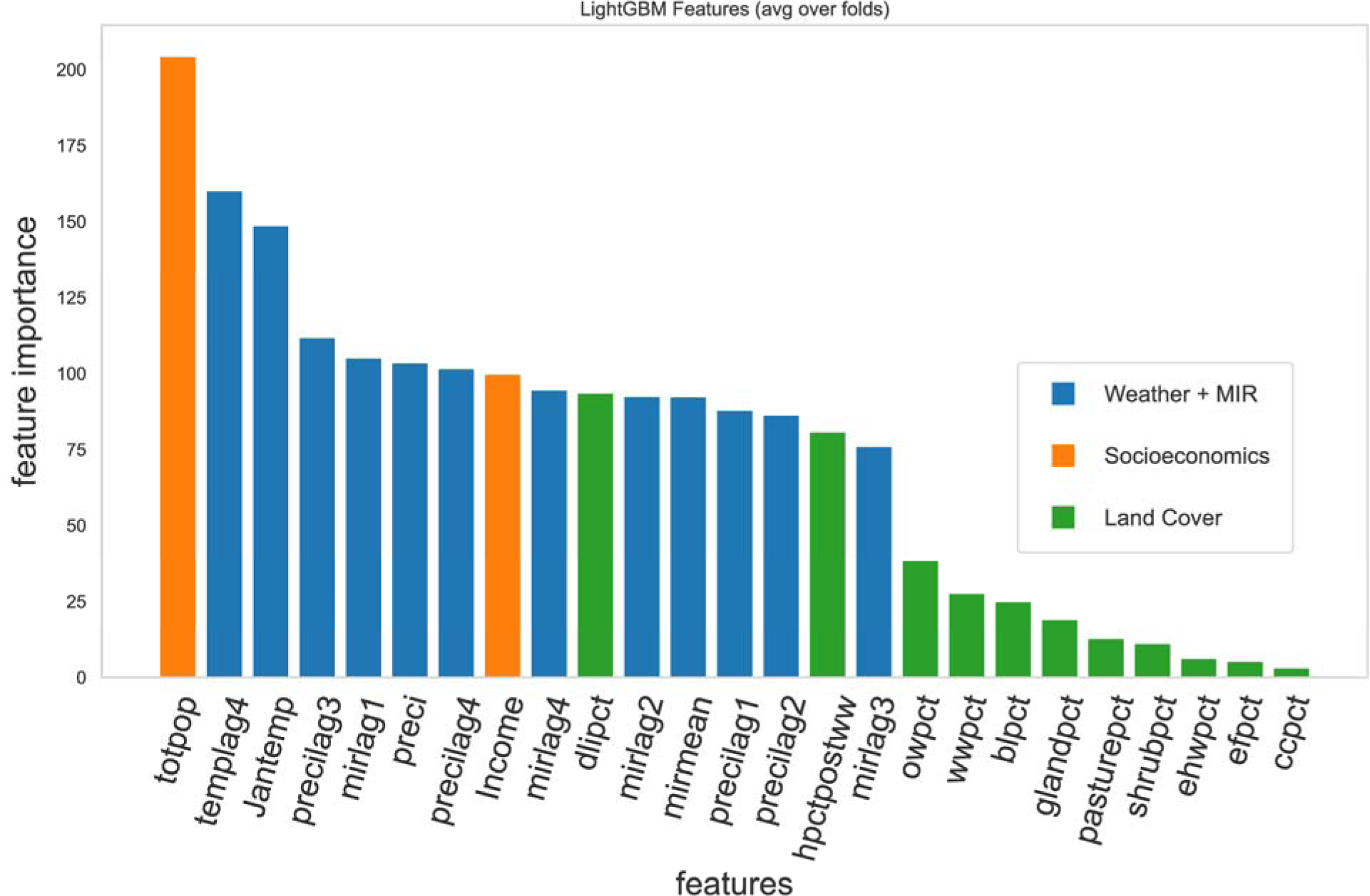
Gini feature importance of the model predicting West Nile Virus cases in the Chicago area, with the 25 variables after removing the highly correlated ones. The higher the y-value, the more important the feature is to the model. The variables are grouped into four main categories, but we have combined the fast-changing weather and MIR into the same category because these variables, as well as the number of cases, are captured with a temporal resolution of one week. The blue bars represent the weekly variables (weather + MIR). Orange bars represent socio-economic variables. The green bars represent the land cover variables. We found that total population is the most important variable in the model. The weekly variables (weather + MIR) are also strong predictors.

**Table 2:**
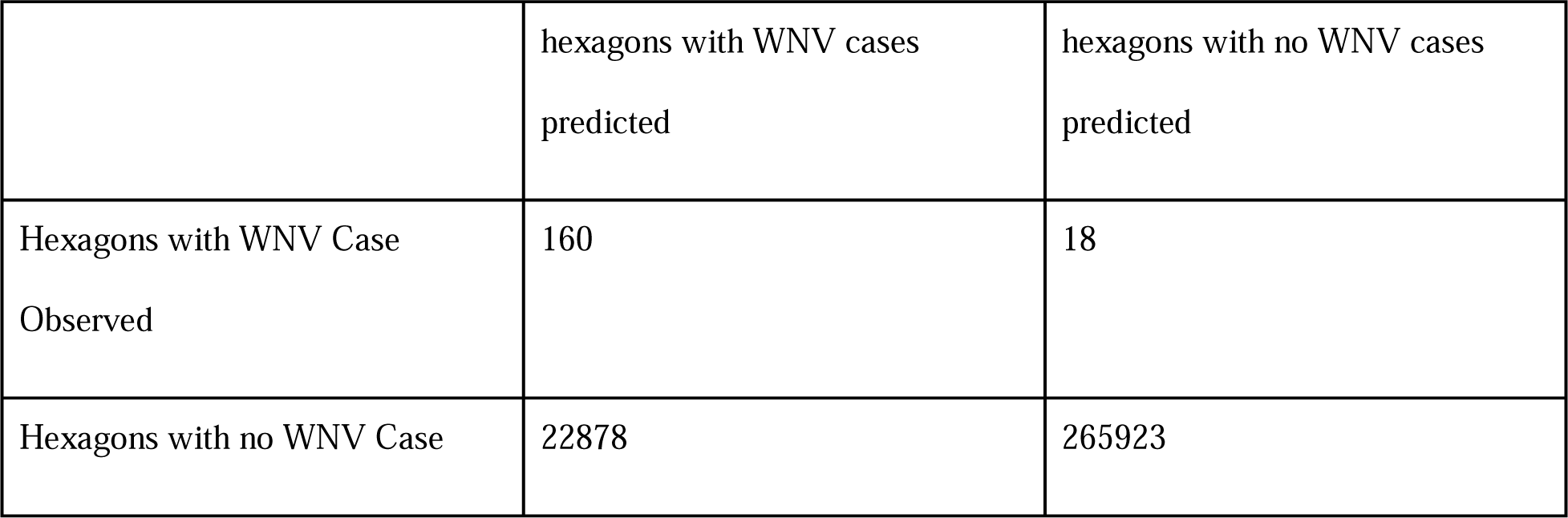
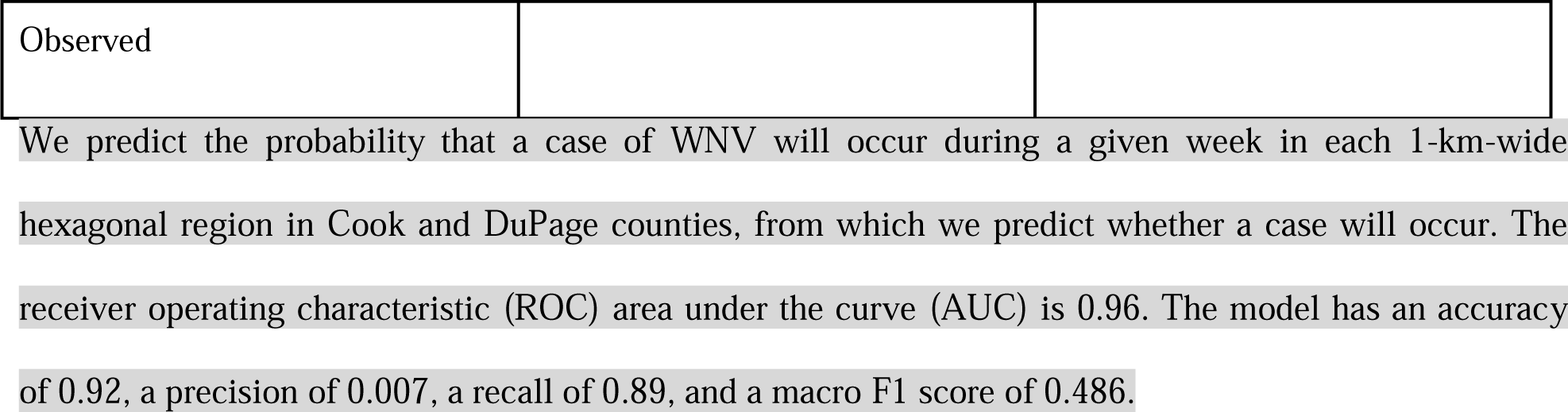
Confusion Matrix of the model including all features.

|  | hexagons with WNV cases<br>predicted | hexagons with no WNV cases<br>predicted |
| --- | --- | --- |
| Hexagons with WNV Case<br>Observed | 160 | 18 |
| Hexagons with no WNV Case | 22878 | 265923 |
| Observed |  |  |

We predict the probability that a case of WNV will occur during a given week in each 1-km-wide hexagonal region in Cook and DuPage counties, from which we predict whether a case will occur. The receiver operating characteristic (ROC) area under the curve (AUC) is 0.96. The model has an accuracy of 0.92, a precision of 0.007, a recall of 0.89, and a macro F1 score of 0.486.

Figure 3 shows that socioeconomic, weather, and mosquito infection factors are candidates for strong predictors. Precipitation variables have relatively low importance among the weather factors, but still have a medium rank among the feature importance. Total population and income level, the two independent socioeconomic variables included in the model, both have high importance in predicting WNV case occurrence. Percentage of housing built after World War II and percentage of low development intensity area are the only strong indicators among the land cover features. We can see a natural gap between MIR 3 weeks before (mirlag3) and percentage of open water (owpct). Therefore, we select the first 16 features for our reduced model.

The cutoff for selecting the features is chosen to maximize the difference between the true positive rate (TPR) and the false positive rate (FPR). Table 2 shows the confusion matrix of the result based on the test set. With the cutoff = 0.625, we obtain a true positive rate (recall or sensitivity) close to 0.89. The precision is about 0.007. This value is not good, but it is still well above the baseline derived from the proportion of positive categories (0.0005) in the dataset. The F1 score is 0.486 and the accuracy is 0.92. Since our model focuses on maximizing recall, this loss in overall performance is to be expected.

### Light GBM model based on reduced features

We re-fit the model using only the features with importance > 20. The feature importance of each predictor in this model is shown in Figure 4, and its performance on the test set is shown in Table 2.

**Fig 4:**
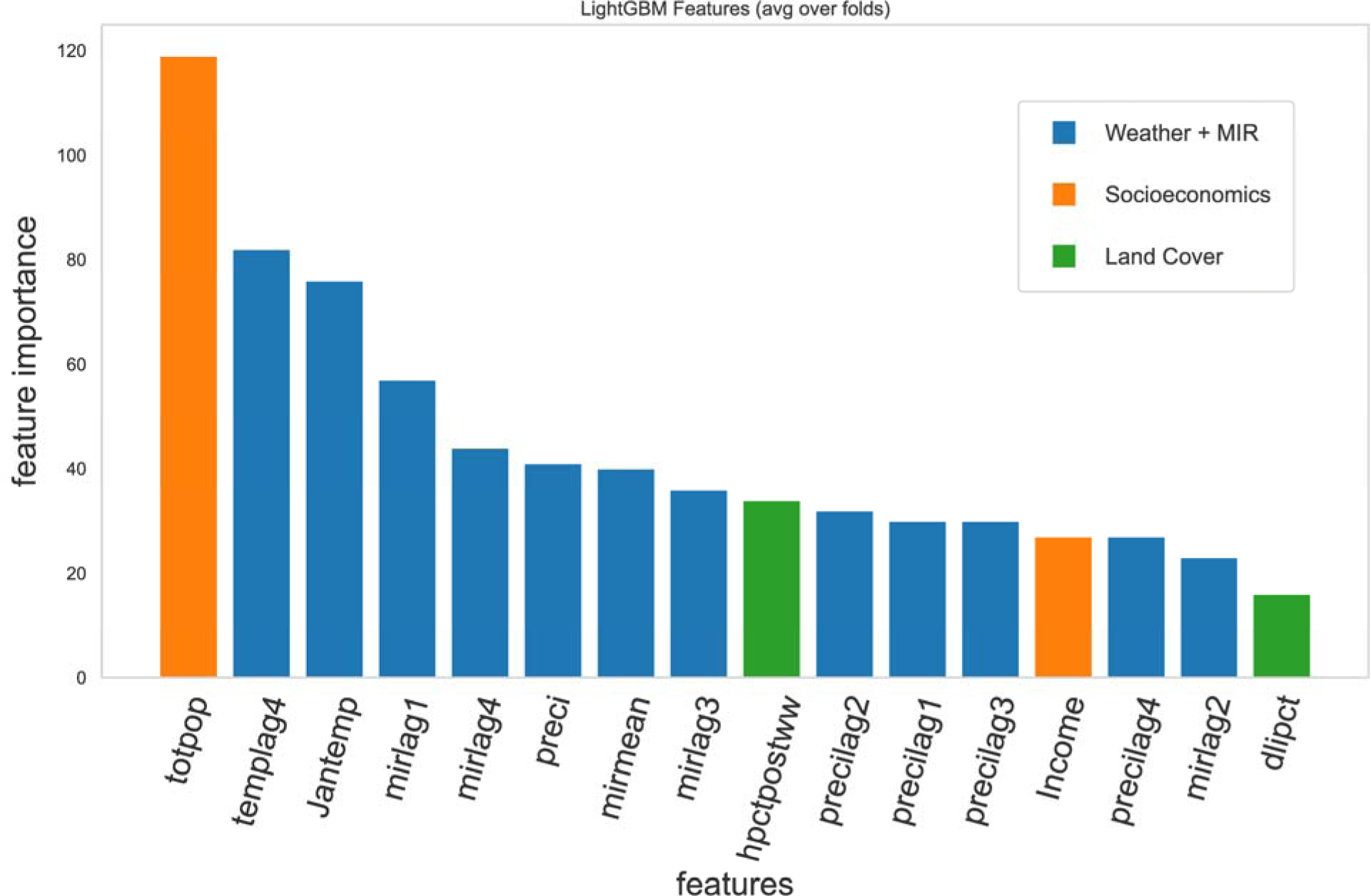
Gini Feature importance of the candidate predictors in the reduced model. Blue, highly dynamic features including weather and mosquito infection rate. Orange: Socioeconomic features. Green, land cover data. The socioeconomic features include total population and income, ranked 1 and 5. The land cover features, share of post-war housing and share of low-intensity development, rank 11 and 15. The most important natural features are the January temperature and the average weekly temperature, followed by the mosquito infection rate. While the ranks may change in individual runs, the feature importance of these factors are close to each other.

The cutoff for selecting the features is chosen to maximize the difference between the true positive rate (TPR) and the false positive rate (FPR). Table 3 shows the confusion matrix of the result based on the test set. With the cutoff = 0.446, we obtain a true positive rate (recall or sensitivity) close to 0.96. The precision is about 0.0034. This value is not good, but it is still well above the baseline derived from the proportion of positive categories (0.0005) in the dataset. The F1 score is 0.45 and the accuracy is 0.83. Since our model focuses on maximizing recall, this loss in overall performance is to be expected.

**Table 3:**
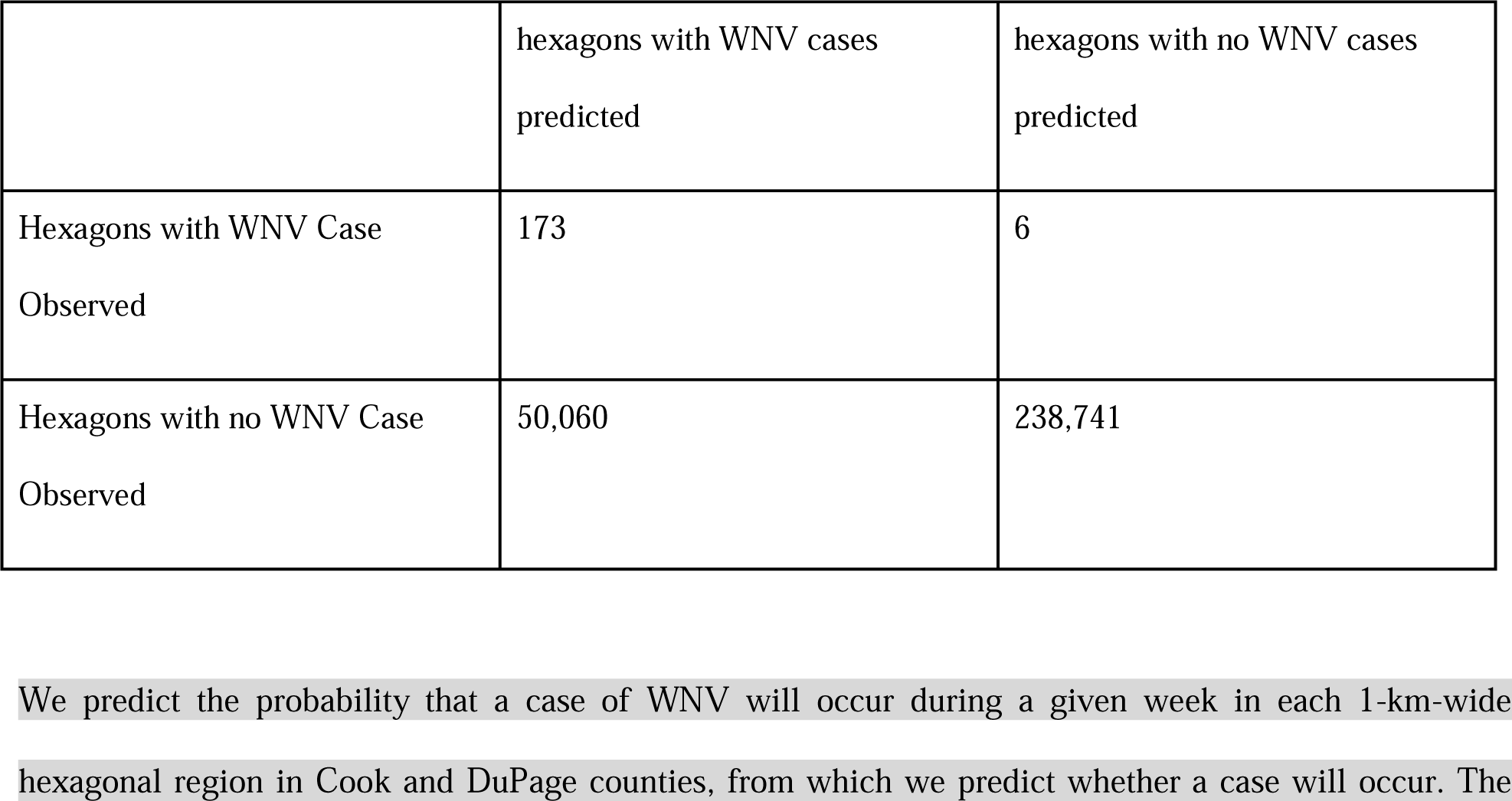
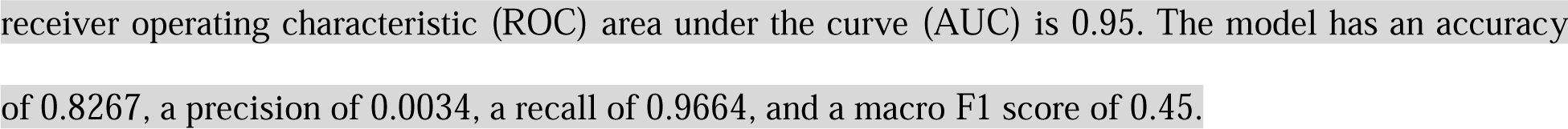
Confusion Matrix of the reduced model.

Based on the above results, we found that the metrics of the reduced model are similar to the model including all 25 low-correlation variables. Therefore, we conclude that the reduced model is sufficient to describe the result.

### Marginal Effects

We examined the marginal effects of all the features by generating partial dependence plots. The slope of the plots shows how much each feature contributes to the model when controlling for the other factors.

Figure 5 shows the partial dependence plot of the factors that predict higher WNV risk as the values of the factors increase. MIR and total population have strong monotonic positive effects. The result is consistent that both disease-carrying mosquitoes and the human population increase the risk of infection. Weekly mean temperature 4 weeks before WNV cases are reported has a strong monotonic positive effect. It is noteworthy that the temperature range is below 30°C, which is approximately the range that promotes mosquito activity and virus replication. January temperature also has a strong monotonic positive effect. A warmer January allows mosquitoes to survive the winter, resulting in larger mosquito populations.

**Fig 5.**
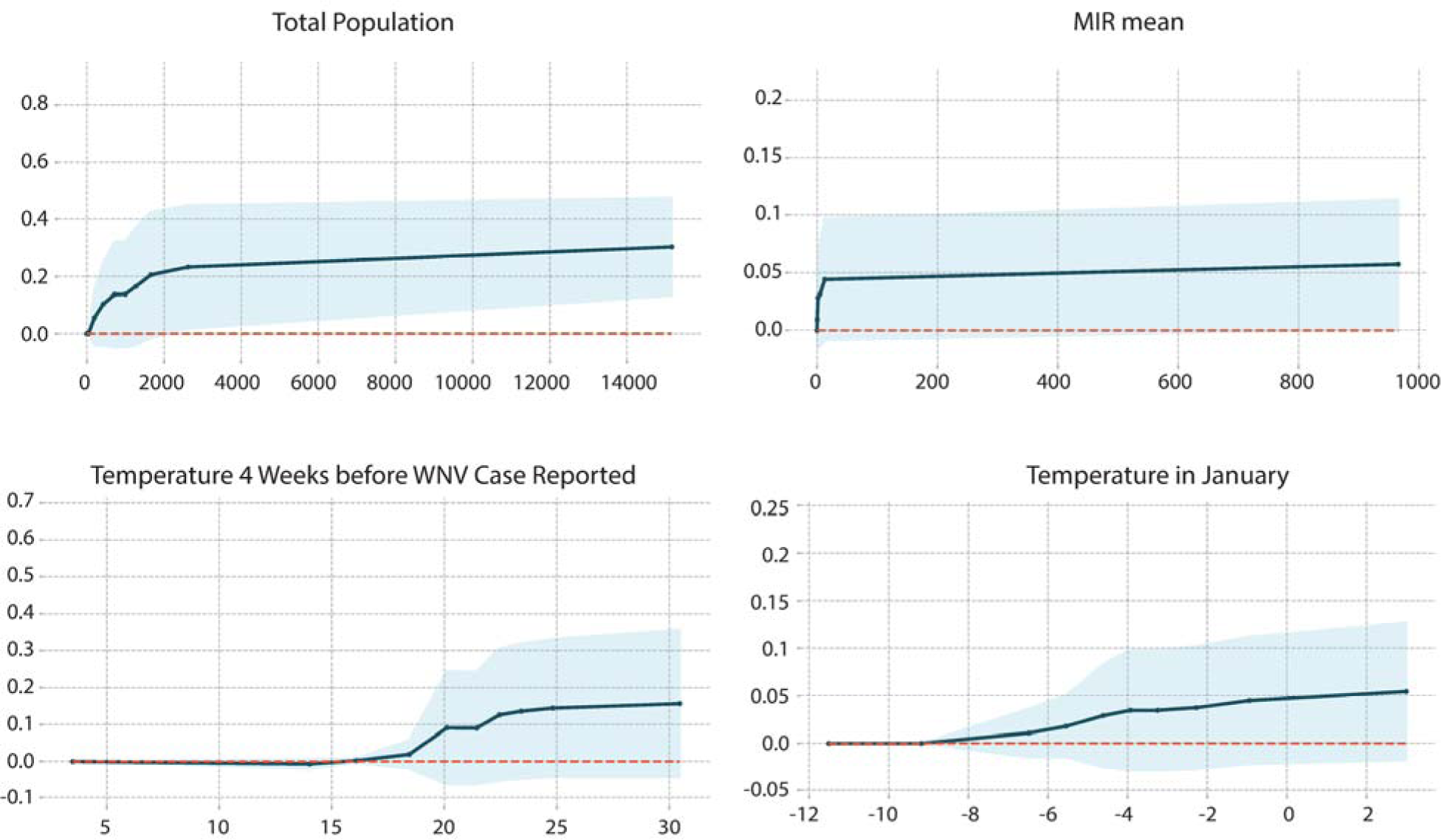
Partial dependence plot of factors with positive effects: total population, mean MIR, temperature 4 weeks before WNV cases are reported, and January temperature. The MIR and theweekly temperatures in 1-4 weeks before also have similar trends as the mean MIR and the temperature of the current week.

On the other hand, as shown in Figure 6, the precipitation variables have non-monotonic effects, i.e., the risk of WNV outbreak first increases and then decreases as precipitation continues to increase. This result is consistent with the existing literature [10,14,26]. While temporary water accumulation provides mosquitoes with more places to lay eggs, excessive precipitation can also wash away mosquito eggs, thus reducing the risk of WNV.

**Fig 6.**
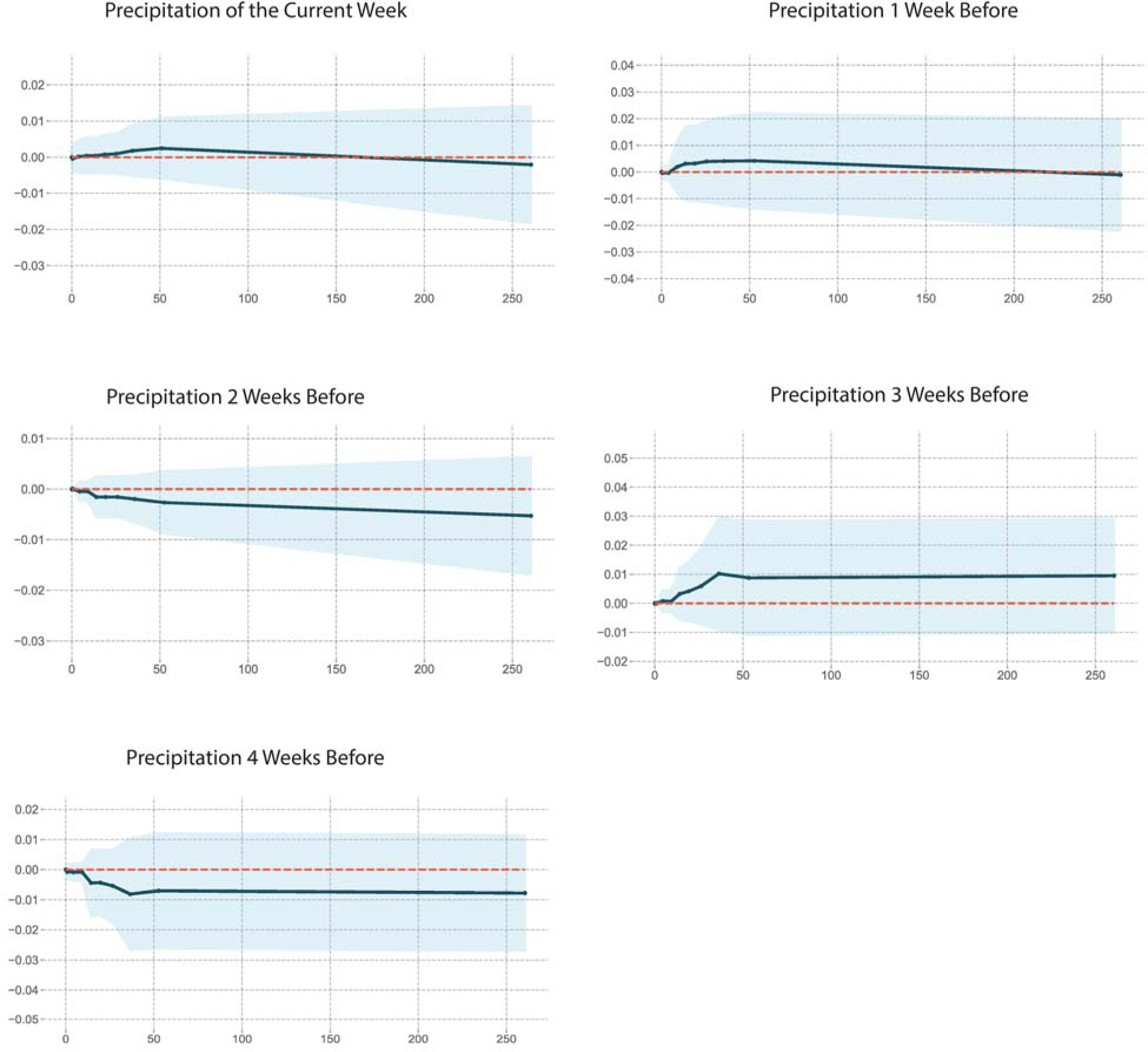
Partial dependence plot of precipitation for the current week and 1-4 weeks prior. Precipitation variables have non-monotonic effects. The marginal effect contributing to WNV risk first increases and then decreases as precipitation increases.

As shown in Figure 7, apart from total population (in Figure 1), land cover and other socioeconomic features have relatively small effects. We don’t observe a marginal effect of income, although it is presented in the feature selection. House age and land development intensity both have small effects on WNV case prediction.

**Fig 7.**
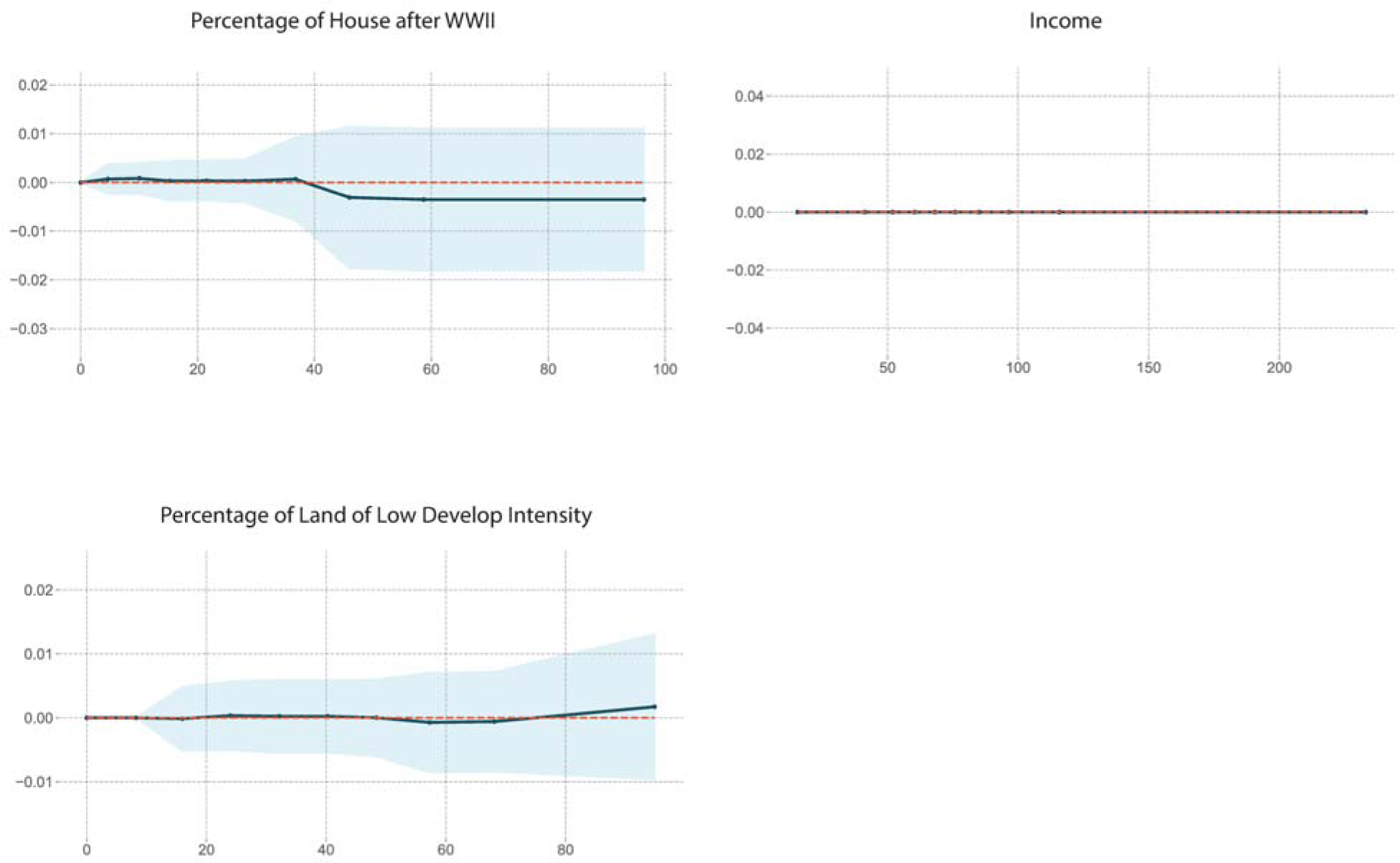
Partial dependence plot of socioeconomics and land cover features. The socioeconomics and land cover features are not very strongly represented. There is no marginal effect of income. The percentage of houses built after World War II has a slight negative effect, indicating that people living in older neighborhoods have higher WNV risks. Meanwhile, the percentage of less developed land has a slight positive effect.

## Conclusion

We performed two-step light GBM procedures to identify a minimum model. We evaluated the ROC-AUC score, accuracy, recall, precision and F-1 score of the models. We found that the reduced model has a worse performance than the linear models of Karki, et. al. [26], while the full model has a similar performance. Therefore, we kept all 25 parameters in the model for prediction. We have found that the natural effects including January temperature, weekly temperature (lagged 0-4 weeks), weekly precipitation (lagged 0-4 weeks), and weekly MIR (lagged 0-4 weeks), as well as the total population are the dominant features that are strongly correlated with the incidence of West Nile virus human cases.

The light GBM model is better at detecting the positive cases, i.e. higher recall. We found consistent features with Karki, et al. that mosquito infection rate, temperature and their lag effects are important factors [26]. This result was further confirmed with PD plots. We also found the behavior of precipitation factors consistent with the literature [10,14,26], being strong predictors with non-monotonic marginal effects. In addition, we found that the percentage of houses built after World War II, which is not included in the original work, is quite important. While income is selected as a predictor by the final model, the PD plot has shown that it has no marginal effects.

The model based only on selected key factors performs similarly to the model that includes all other factors. In addition, both the number of cases and the weather vary on a weekly basis, while the land cover and socioeconomic data are static. Therefore, the effect of the socioeconomic characteristics could be masked by the correlated characteristics of lagged MIR and lagged temperature.

One concern was that the behavior of the model may differ by the income of the area, as income disparities may affect diagnosis rates, surveillance efforts, and distribution of land cover and housing variables. Therefore, the light GBM model fitting was repeated for subsets of the data consisting of the areas with above-median income and the areas with below-median income (S3). These stratified models were similar to each other and to the full model, indicating that the predictive capabilities of this model are not predicated on income groupings.

In conclusion, our light GBM model provides an alternative way to predict the probability of an area having a WNV case or not. The performance in terms of ROC-AUC is very close to the previous work [26] and is much better at detecting the area where there is actually a case. We also have a clearer relationship between temperature and precipitation, mosquito infection, and West Nile virus. In addition, we identified weak effects of socioeconomics and land cover. The risk of contracting WNV does not appear to be related to income in these data. However, other factors may relate to income and WNV detection that are not possible to study with these data, such as variation in diagnosis rates.

The results of this study can be used as a guideline to develop a threshold for public health intervention.

## Supporting information

description for area conversion

Modeling with Socioeconomic Data with 5-year rolling average 2010-2017

Models stratified by income.

## Data Availability

All data produced in the present study are available upon reasonable request to the authors

## Acknowledgements

The authors thank the NCSA Center-Directed Discretionary Research (CDDR) for funding this project. The authors would like to thank the SPIN program at NCSA for supporting the student who is the first author of the paper. The authors would like to thank the HAL cluster and support team for providing the computational resources to complete the work. The author would also like to acknowledge the efforts of the NCSA Industry Group for supporting the work. The authors would like to thank Dr. Christina Fliege for her editorial suggestions on this manuscript. The authors would like to thank Mr. Mingyu Yang for his help in retrieving and preprocessing the census data.

