## Supplementary material for "Two-Step Light Gradient Boosted Model to identify human West Nile Virus infection risk factor in Chicago": description for area conversion

### S1. Socioeconomic data cleaning and transformation

The socioeconomic columns in Karki et al. are approximated by the 2016 values for all years. To investigate whether the more recent socioeconomic data could make the model more accurate, we obtained socioeconomic data from the US Census Bureau [[33]](https://paperpile.com/c/FpOHrF/5wsw) with the granularity of census tracts. We requested a 5-year average of the data from 2010 to 2017. The number of rows was consistent because the census tract did not change during the time period. The column numbers are not consistent. We filtered and cleaned the dataset using the following steps.

First, we dropped all empty columns whose values are marked with an "X". Then we dropped the columns with the "margin of error" of the variables to simplify the problem. Then we dropped the extra columns that do not span all the years of interest. Some columns describe the same type of variable, but are named differently. For example, in 2010 one of the variables was called "Estimate; SEX AND AGE - Male". On the other hand, in 2013, the same variable was called "Estimate; SEX AND AGE - Total Population - Male". Therefore, we implemented a cosine similarity metric to standardize these column names. This method is used to measure or calculate how similar the set or documents are, regardless of size. Mathematically, it measures the cosine of the angle between two vectors projected in multidimensional space. The values of cosine range from 0 to 1. The closer the value is to 1, the higher the probability that two sentences have similar words or represent similar things. After merging the columns, we evaluated the missing values. We dropped all columns with more than 10% missing values. For the remaining columns, we applied a mean imputation for the rest of the columns.

Finally, we removed all highly collinear independent variables. We calculated the Pearson correlation between the independent variables. If two variables have a correlation higher than 0.8, we drop one of them.

The data are available at different spatial resolutions. The WNV cases and the mosquito infection rate are in units of hexagons with 1 km on each side. We obtained the shapefiles overlapping hexagons and census tracts from Karki et al. and assumed that the socioeconomic data are homogeneously distributed within each census tract.

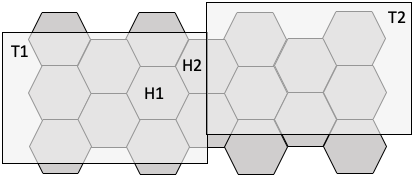

**S1 Figure. 1. Census tract overlapping with hexagons.** Hexagon H1 is completely covered by census tract T1. Hexagon H2 overlaps census tracts T1 and T2. In H1, the values of intensive properties are the same as in T1. The value of the extensive properties is proportional to the area. In H2, the values of the properties in the overlapping areas with T1 and T2 are calculated separately, and then a weighted average is taken.

In S1. Fig. 1, hexagon H1 overlaps only with census tract T1, and hexagon H2 overlaps with census tracts T1 and T2. The conversion methods are different for extensive and intensive tract variables. The extensive properties, if homogeneous, are proportional to area, while the intensive properties remain the same as area changes. For example, population is an extensive property, while population density is an intensive property. If a hexagon is completely covered by a census tract (e.g., H1), the conversion is as follows. For the extensive properties, it would be $V_{h}=\frac{A_{h}}{A_{t}}\times V_{t}$, where $A_{h}$ is the area of the hexagon, $A_{t}$ is the area of the census tract, $V_{h}$ is the value of the property in the hexagon, and $V_{t}$ is the value of the extensive property in the census tract. For the intensive properties, it would be $V_{h}=V_{t}$. If the hexagon overlaps multiple census tracts, we take a weighted average by the overlapping areas.

For the intensive properties, the value $V_{hi}$ n the areas overlapping census tract $T_{i}$ is equal to $V_{ti}$. The hexagon value $V_{h}$ is the weighted average:

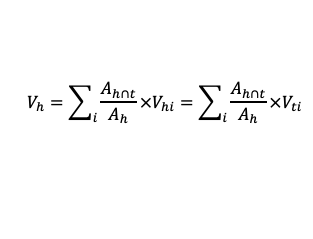
 (Eq.1)

For the extensive properties, we first convert the value $V_{i}$ to the intensive property $V_{i}/A_{i}$ for both the hexagon and the census tract, and then plug it into Eq.1. We have:

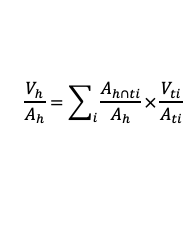
 (Eq.2)

Canceling $A_{h}$, we reach:

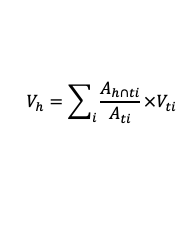
(Eq.3)

where	$A_{h}$= hexagon area, in $m^{2}$for our data.

$A_{t}$= census tract area, in $m^{2}$for our data

$V_{t}$= the value of extensive property at the census tract level

$V_{h}$= the value of extensive property at the hexagon level.

i = the index of each hexagon portion overlapping with a different census tract.

In summary, the equation calculates the **"sum of all values for a given hexagon h".**
