## Supplementary material for "Two-Step Light Gradient Boosted Model to identify human West Nile Virus infection risk factor in Chicago": Modeling with Socioeconomic Data with 5-year rolling average 2010-2017

### S2. Modeling with Socioeconomic Data with 5-year rolling average 2010-2017

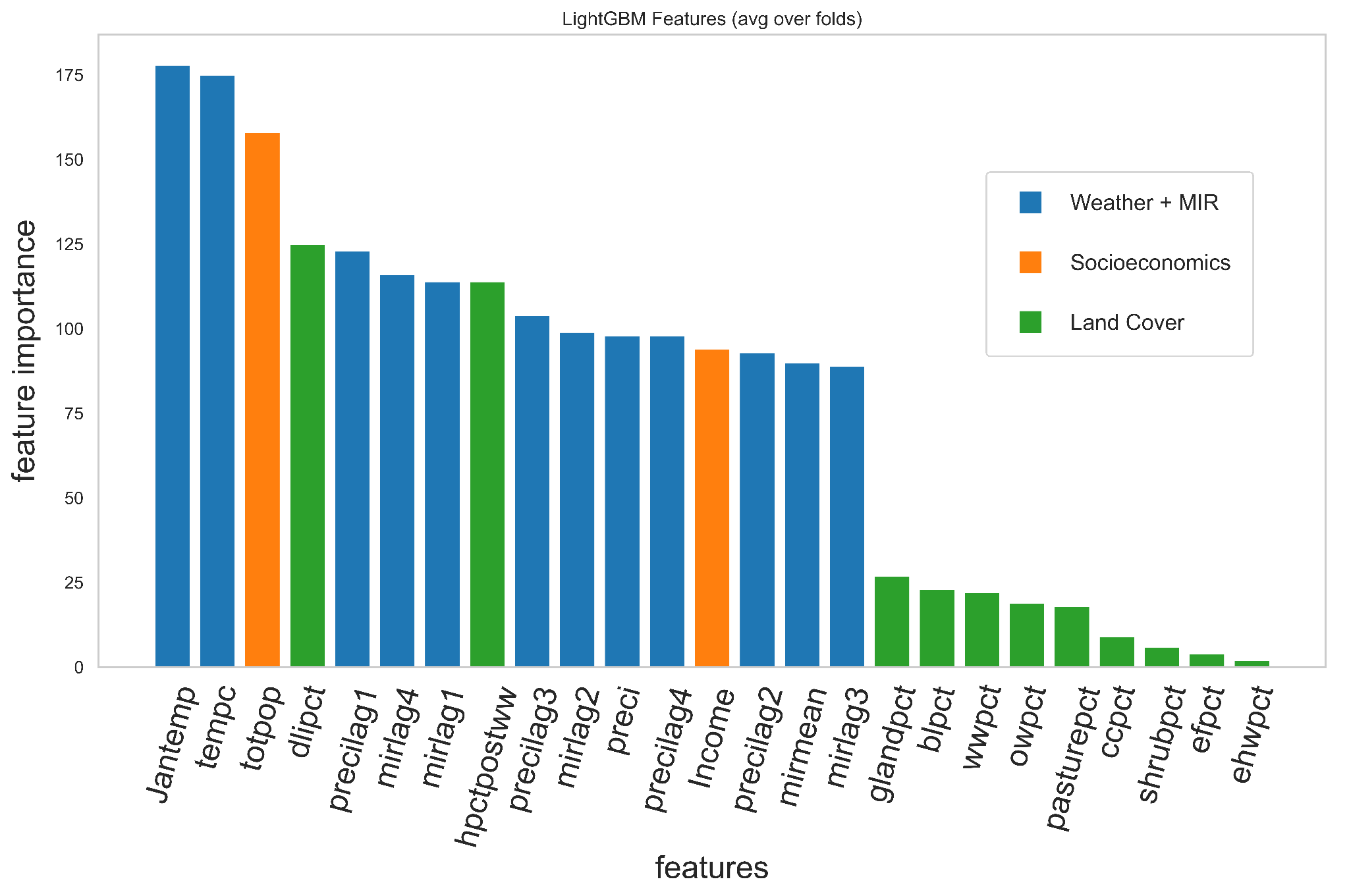

**S2 Figure 1: Gini Feature importance of the model predicting the cases of West Nile Virus in the Chicago area, based on the weather and landscape data from Kark et al. (2020), and 5-year rolling average socioeconomic data from the US Census Bureau, i.e. varied socioeconomic data.** The higher the y-value, the more important the feature is to the model. The variables are grouped into four main categories, but we have combined the fast-changing weather and MIR into the same category because these variables, as well as the number of cases, are captured with a temporal resolution of one week. The blue bars represent the weekly variables (weather + MIR). Orange bars represent socio-economic variables. The green bars represent the land cover variables. We found that total population is the most important variable in the model. The weekly variables (weather + MIR) are also strong predictors.

|  | hexagons with WNV cases predicted | hexagons with no WNV cases predicted |
| --- | --- | --- |
| Hexagons with WNV Case Observed | 172 | 7 |
| Hexagons with no WNV Case Observed | 61094 | 227707 |

**S2 Table 1:**  **Confusion Matrix of the model based on the data from Kark et al. (2020)**[[26]](https://paperpile.com/c/FpOHrF/iycs)**, i.e. fixed socioeconomic data. Same as Table 1 in the Results part.** We predict the probability that a case of WNV will occur during a given week in each 1-km-wide hexagonal region in Cook and DuPage counties, from which we predict whether a case will occur. The receiver operating characteristic (ROC) area under the curve (AUC) is 0.93. The model has an accuracy of 0.79, a precision of 0.0028, a recall of 0.96, and a macro F1 score of 0.3697.

|  | hexagons with WNV cases predicted | hexagons with no WNV cases predicted |
| --- | --- | --- |
| Hexagons with WNV Case Observed | 75 | 9 |
| Hexagons with no WNV Case Observed | 40,972 | 117,503 |

**S2 Table 2:**  **Confusion Matrix of the model based on the weather and landscape data from Kark et al. (2020)** [[26]](https://paperpile.com/c/FpOHrF/iycs)**, and 5-year rolling average socioeconomic data from the US Census Bureau, i.e. varied socioeconomic data.** We predict the probability that a case of WNV will occur during a given week in each 1-km-wide hexagonal region in Cook and DuPage counties, from which we predict whether a case will occur. The receiver operating characteristic (ROC) area under the curve (AUC) is 0.895. The model has an accuracy of 0.7415, a precision of 0.0018, a recall of 0.8929, and a macro F1 score of 0.4276.

We obtained the 5-year rolling average of 2010-2017 socioeconomic data from the US Census Bureau and mapped it to hexagons. We build a lightweight GBM model based on the data and compare it with the model built on 2016 socioeconomic data used by Karki et al. [[26]](https://paperpile.com/c/FpOHrF/iycs). We found that the top features are quite similar. However, since the data set is smaller, the performance is worse. Therefore, to compare the results with Karki et al. [[26]](https://paperpile.com/c/FpOHrF/iycs), we decide to use the 2016 socioeconomics data instead of the 5-year rolling average.
