## Supplementary material for "Two-Step Light Gradient Boosted Model to identify human West Nile Virus infection risk factor in Chicago": Models stratified by income.

### S3. Models stratified by income.

We also investigate whether high-income and low-income groups have different predictive factors. Therefore, we performed a light GBM analysis on the data stratified by income. The high-income dataset is the dataset containing hexagons with average annual household income >= $56,000, and the rest of the data is the low-income dataset. We have plotted the feature importance in S3. Fig. 1, and the confusion matrices are in S3 Tables 1 and 2. We found that the features selected in the high- and low-income models are similar. Therefore, both high- and low-income groups can be described by the same models.

a)
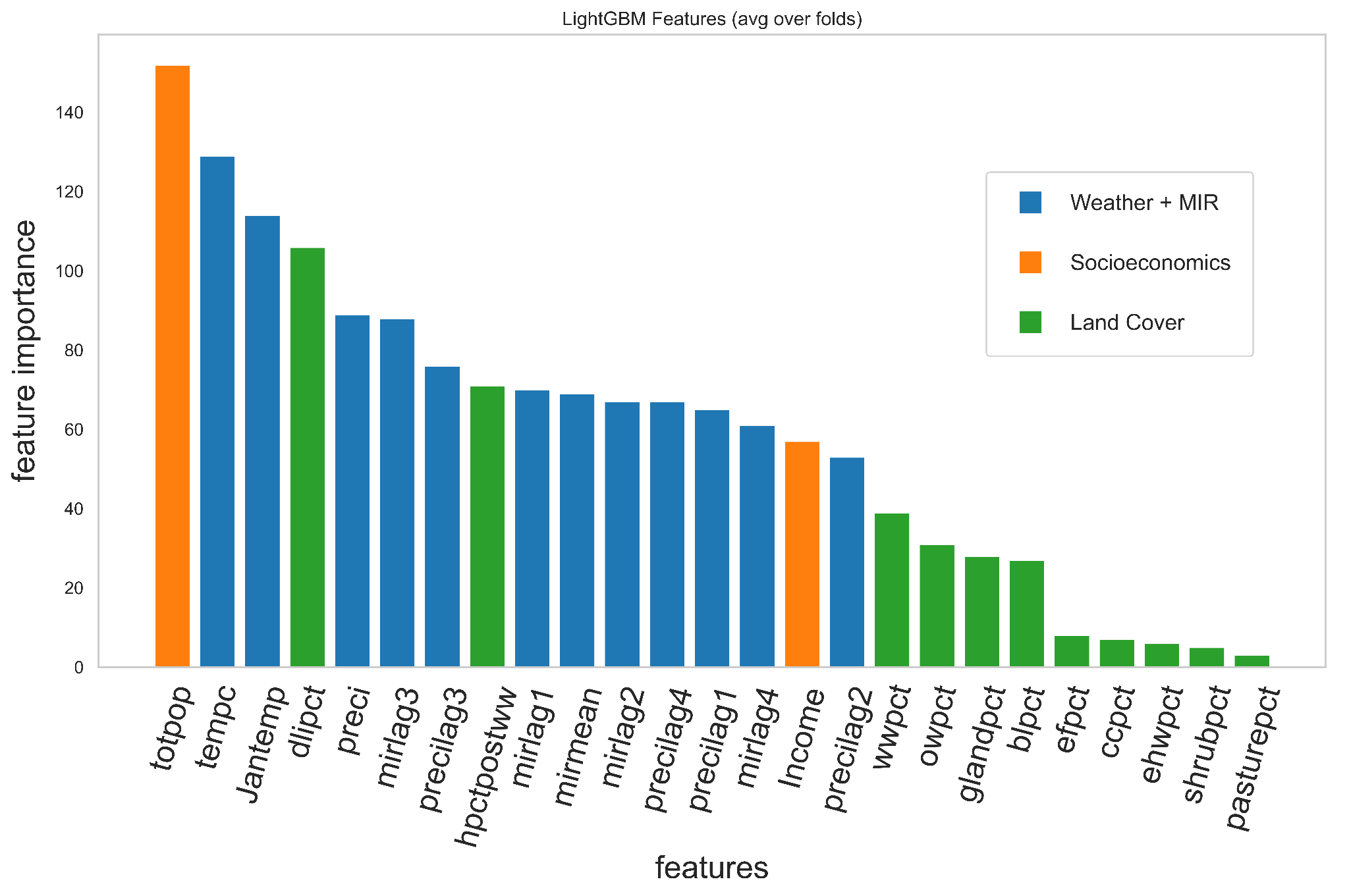

b)
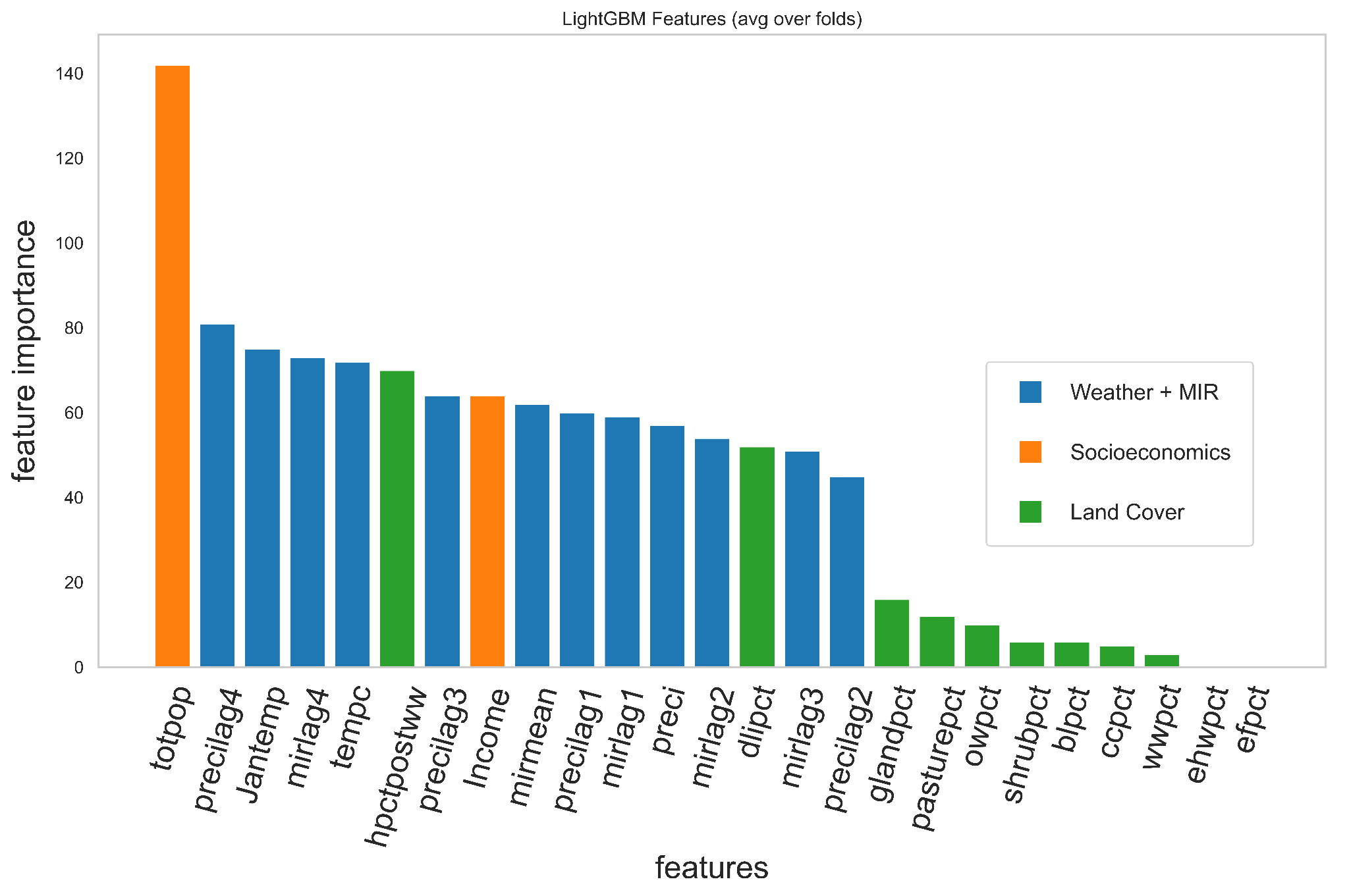

**S3 Figure 1. Feature importance based on income-stratified data.** Blue, highly dynamic features such as weather and mosquito infection rate. Orange: Socioeconomic features. Green, land cover data. a) High income data (income > $56,000) b) Low income data (income < $56,000). For both models, total population, weather, and mosquito infection rate are the most important factors. The most important land cover characteristics in the model are the percentage of light development intensity and the percentage of houses built after World War II.

|  | hexagons with WNV cases predicted | hexagons with no WNV cases predicted |
| --- | --- | --- |
| Hexagons with WNV Case Observed | 85 | 44 |
| Hexagons with no WNV Case Observed | 13,707 | 195,944 |

**S3 Table 1:**  **Confusion Matrix of the model based on the high-income data.** We predicted the probability of a WNV case occurring during a given week in each 1-km-wide hexagonal region in Cook and DuPage counties, from which we predicted whether a case would occur. The prediction was based on income data greater than or equal to the median income in the Chicago area. The receiver operating characteristic (ROC) area under the curve (AUC) is 0.92. The model has a precision of 0.8694. At a cut-off of 0.6922, the model has a precision of 0.0062, a recall of 0.6589, and a macro F1 score of 0.4892.

|  | hexagons with WNV cases predicted | hexagons with no WNV cases predicted |
| --- | --- | --- |
| Hexagons with WNV Case Observed | 44 | 6 |
| Hexagons with no WNV Case Observed | 20,459 | 58,691 |

**S3 Table 2:**  **Confusion Matrix of the model based on the low-income data.** We predicted the probability of a WNV case occurring during a given week in each 1-km-wide hexagonal region in Cook and DuPage counties, from which we predicted whether a case would occur. The prediction was based on income data below the median income for the Chicago area. The receiver operating characteristic (ROC) area under the curve (AUC) is 0.92. The model has a precision of 0.8763. At a cut-off of 0.0708, the model has a precision of 0.0021, a recall of 0.88 and a macro F1 score of 0.4279.
